# Risk Factors for COVID-19-associated hospitalization: COVID-19-Associated Hospitalization Surveillance Network and Behavioral Risk Factor Surveillance System

**DOI:** 10.1101/2020.07.27.20161810

**Authors:** Jean Y. Ko, Melissa L. Danielson, Machell Town, Gordana Derado, Kurt J. Greenlund, Pam Daily Kirley, Nisha B. Alden, Kimberly Yousey-Hindes, Evan J. Anderson, Patricia A. Ryan, Sue Kim, Ruth Lynfield, Salina M. Torres, Grant R. Barney, Nancy M. Bennett, Melissa Sutton, H. Keipp Talbot, Mary Hill, Aron J. Hall, Alicia M. Fry, Shikha Garg, Lindsay Kim, COVID-NET Investigation Group

## Abstract

**Background:** Identification of risk factors for COVID-19-associated hospitalization is needed to guide prevention and clinical care.

**Objective:** To examine if age, sex, race/ethnicity, and underlying medical conditions is independently associated with COVID-19-associated hospitalizations.

**Design:** Cross-sectional.

**Setting:** 70 counties within 12 states participating in the Coronavirus Disease 2019-Associated Hospitalization Surveillance Network (COVID-NET) and a population-based sample of non-hospitalized adults residing in the COVID-NET catchment area from the Behavioral Risk Factor Surveillance System.

**Participants:** U.S. community-dwelling adults (≥18 years) with laboratory-confirmed COVID-19-associated hospitalizations, March 1- June 23, 2020.

**Measurements:** Adjusted rate ratios (aRR) of hospitalization by age, sex, race/ethnicity and underlying medical conditions (hypertension, coronary artery disease, history of stroke, diabetes, obesity [BMI ≥30 kg/m^2^], severe obesity [BMI≥40 kg/m^2^], chronic kidney disease, asthma, and chronic obstructive pulmonary disease).

**Results:** Our sample included 5,416 adults with COVID-19-associated hospitalizations. Adults with (versus without) severe obesity (aRR:4.4; 95%CI: 3.4, 5.7), chronic kidney disease (aRR:4.0; 95%CI: 3.0, 5.2), diabetes (aRR:3.2; 95%CI: 2.5, 4.1), obesity (aRR:2.9; 95%CI: 2.3, 3.5), hypertension (aRR:2.8; 95%CI: 2.3, 3.4), and asthma (aRR:1.4; 95%CI: 1.1, 1.7) had higher rates of hospitalization, after adjusting for age, sex, and race/ethnicity. In models adjusting for the presence of an individual underlying medical condition, higher hospitalization rates were observed for adults ≥65 years, 45-64 years (versus 18-44 years), males (versus females), and non-Hispanic black and other race/ethnicities (versus non-Hispanic whites).

**Limitations:** Interim analysis limited to hospitalizations with underlying medical condition data.

**Conclusion:** Our findings elucidate groups with higher hospitalization risk that may benefit from targeted preventive and therapeutic interventions.

## INTRODUCTION

As of June 26, 2020, over 9 million cases of Coronavirus Disease 2019 (COVID-19), the disease caused by SARS-CoV-2, have been reported worldwide (1); over 2 million cases, including >120,000 deaths, have been reported in the United States (2). Older age and underlying medical conditions are prevalent among cases (3, 4, 5, 6, 7, 8, 9, 10, 11, 12, 13). Based on preliminary estimates, approximately 30% of U.S. laboratory-confirmed COVID-19 cases were among adults aged ≥65 years (7, 8) and about one third had underlying medical conditions (9). Among U.S. hospitalized cases, diabetes mellitus (8, 9, 10, 11, 12, 13, 14), hypertension (10, 11, 12, 13, 14), cardiovascular disease (8, 9, 10, 14) obesity (10, 11, 13, 14), and chronic lung disease (8, 9, 10) were common. However, the risk of hospitalization imparted by underlying medical conditions is not clear; many of these conditions, e.g., obesity (15), hypertension (16), and diabetes (17), are also prevalent in the general U.S. population.

Similarly, the risk of hospitalization related to sex and race/ethnicity is unclear. An estimated 60% of New York patients hospitalized for COVID-19 were male (11); however, other studies have found the male-female distribution among COVID-19 hospitalizations to be similar to the general U.S. population (50%) (10,18). Non-Hispanic black adults comprised a greater proportion of hospitalized COVID-19 cases compared to the community population in 14 states (10) and to overall hospitalizations in Georgia (18).

Two studies of communities served by single health care systems in Louisiana (19) and in New York City and Long Island (20) assessed the independent risk for hospitalization among adults who tested positive for SARS-CoV-2 (19, 20); however these studies did not account for the underlying distribution of age, sex, race/ethnicity and underlying medical conditions in these communities.

To better understand the independent association of age, sex, race/ethnicity, and underlying medical conditions with COVID-19-associated hospitalization relative to the non-hospitalized community-dwelling population, we calculated rate ratios for adults with and without select underlying medical conditions, adjusted for age, sex, and race/ethnicity, using data from the Coronavirus Disease 2019-Associated Hospitalization Surveillance Network (COVID-NET) and the Behavioral Risk Factor Surveillance System (BRFSS), two large multi-state surveillance systems.

## METHODS

### Surveillance data sources and definition of cases

COVID-NET is an all age population-based surveillance system of laboratory-confirmed COVID-19-associated hospitalizations. To be included as a case, patients must have a positive SARS-CoV-2 test no more than 14 days before admission or during hospitalization; be a resident of the pre-identified surveillance catchment area; and be admitted to a hospital where residents of the surveillance catchment area receive care. Medical chart abstractions using a standard case report form are performed by trained surveillance officers to collect additional data such as patient demographics, underlying medical conditions, clinical course, and outcomes data. Additional COVID-NET details are described elsewhere (10, 21). This study includes 70 counties in 12 participating states (California, Colorado, Connecticut, Georgia, Maryland, Michigan, Minnesota, New Mexico, New York, Oregon, Tennessee, and Utah).

The BRFSS is a nationwide cross-sectional telephone survey that collects state-based data on health-related risk behaviors, chronic health conditions, and use of preventive services from more than 400,000 community-dwelling adults (≥18 years) each year (22). The BRFSS was used to provide estimates of the non-hospitalized population in the 70 COVID-NET counties included in this study, herein referred to as the COVID-NET catchment area. The percentage of adults with the select underlying medical conditions of interest by demographic subgroup (sex, age group, race/ethnicity group) were calculated from BRFSS respondents residing in the COVID-NET catchment area. These responses were then weighted to the total population at risk for hospitalization residing in the catchment area using an iterative proportional fitting method, which includes categories of age by gender, race and ethnicity groups, education levels, marital status, regions within states, gender by race and ethnicity, telephone source, renter or owner status, and age groups by race and ethnicity to improve the degree and extent to which the BRFSS sample properly reflects the sociodemographic make-up of our geographic area of interest (22). Weights also accounted for survey design, probability of selection, nonresponse bias, and non-coverage error (22). To understand if the prevalence of underlying medical conditions in the COVID-NET catchment area was different from national estimates, nationwide BRFSS data were used. All weighted population estimates were calculated using 2018 BRFSS data for each characteristic and underlying medical condition except hypertension; 2017 was the most recent year of available BRFSS data that included hypertension questions.

To match the population captured by BRFSS, this analysis was restricted to community-dwelling adults (≥ 18 years) residing in the 70 COVID-NET counties in 12 states with available data on underlying medical conditions (Figure 1). As of June 23, 2020, there were a total of 5,715 adult COVID-19-associated hospitalizations eligible for inclusion in our analysis; 5,416 adults had underlying medical condition data and composed the analytic population for this study.

**Figure 1.**
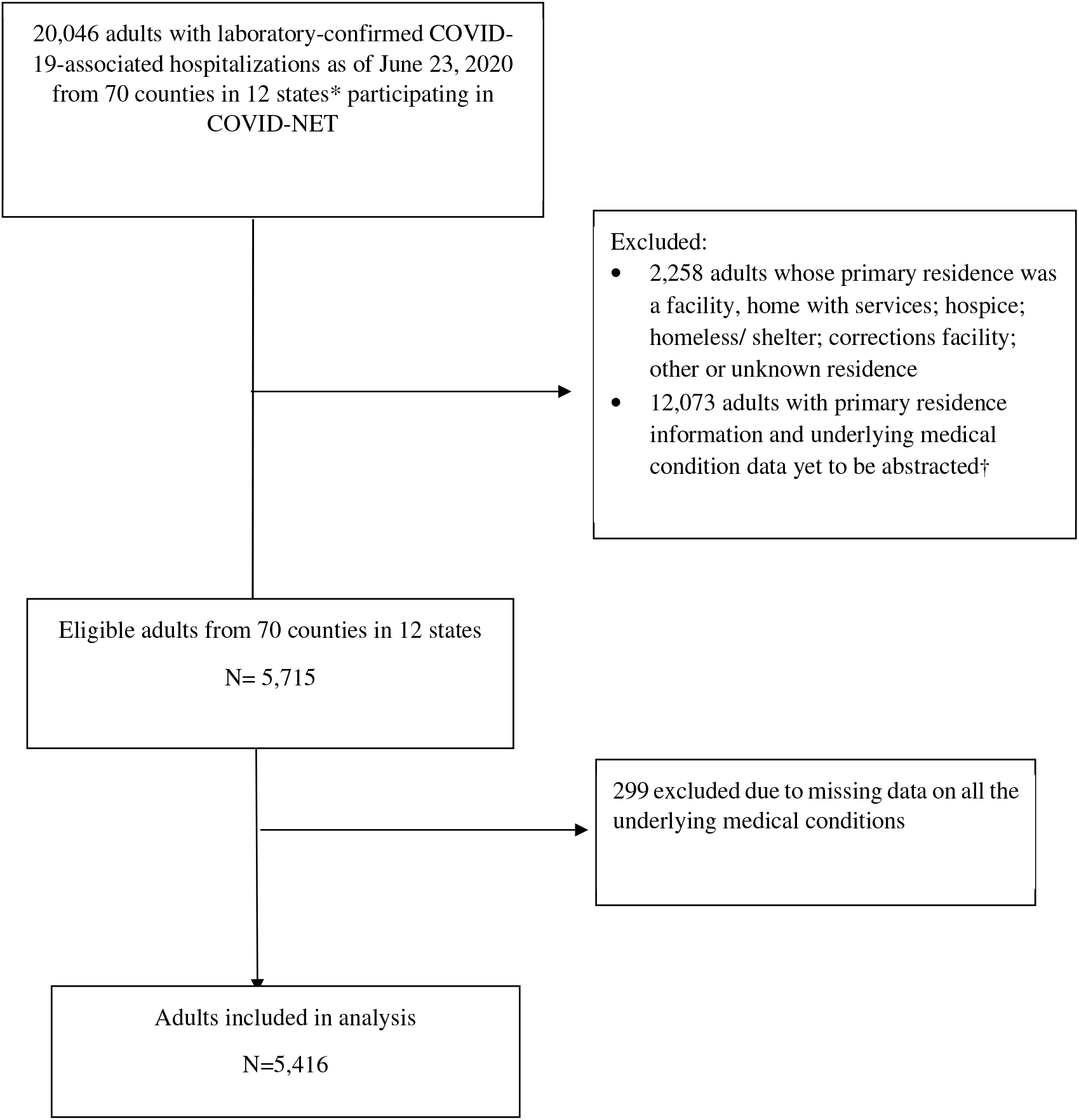
Analytic Population Flow Diagram, Coronavirus Disease 2019-Associated Hospitalization Surveillance Network, March 1-June 23, 2020. *California, Colorado, Connecticut, Georgia, Maryland, Michigan, Minnesota, New Mexico, New York, Oregon, Tennessee, and Utah. †Additional data beyond the minimum required data elements (Case Identification Number, state, case type [pediatric vs. adult], age, admission date, sex, and SARS-CoV-2 test result [test type, test date, test result) to calculate age-stratified COVID-19-associated hospitalization rates may be subject to a time lag for submission to CDC.

### Variable definitions

In COVID-NET, the presence of underlying medical conditions was ascertained if the condition (hypertension; history of myocardial infarction, coronary artery disease, coronary artery bypass grafting; stroke; diabetes mellitus; chronic kidney disease; asthma; chronic obstructive pulmonary disease [COPD]) was present in the patient’s medical chart that detailed their COVID-19-associated hospitalization. In BRFSS, underlying medical conditions were based on self-report to the question: “Has a doctor, nurse, or other health professional ever told you that you had… “(high blood pressure; heart attack also called myocardial infarction, angina or coronary heart disease; stroke; diabetes, chronic kidney disease; asthma; COPD, emphysema, or chronic bronchitis).

Histories of myocardial infarction, coronary artery disease, and coronary artery bypass grafting (only available in COVID-NET) were categorized as coronary artery disease. In BRFSS, adults who self-reported having high blood pressure and answered “yes” to the subsequent question “are you currently taking medication for your high blood pressure?” were categorized as having hypertension. In COVID-NET data, body mass index (BMI) was calculated using height and weight listed in medical charts; if these data were not available, recorded BMI was used. In BRFSS, self-reported height and weight were used to calculate BMI. BMI was then categorized as obese (≥30 kg/m^2^) or severely obese (≥40 kg/m^2^).

For both COVID-NET and BRFSS data, we created an “any condition” variable (which included presence of history of coronary artery disease; stroke; diabetes; obesity; severe obesity; chronic kidney disease; asthma; COPD) and “number of conditions” variable (0; 1; 2; 3+). Hypertension was not included in the “any condition” or “number of conditions” variables because COVID-NET catchment estimates for hypertension were derived from 2017 BRFSS estimates and could not be integrated with the other 2018 estimates of underlying medical conditions. Although hypertension is not included in these composite variables, in 2017, 14% of adults with treated hypertension also had at least one other underlying medical condition examined in this analysis. Additional details are available in Supplemental Table 1. The following categories were defined for age (18-44; 45-64; ≥65 years), sex (male; female), and race/ethnicity (non-Hispanic white; non-Hispanic black; other). Other races and ethnicities besides non-Hispanic white and non-Hispanic were aggregated due to small cell sizes from the COVID-NET catchment area once these data were stratified by age, sex and underlying medical conditions.

### Statistical analysis

Demographic characteristics were tabulated among hospitalized COVID-19 cases overall and by underlying medical condition. The prevalence of select underlying medical conditions was calculated among COVID-19-associated hospitalizations, the COVID-NET catchment area, and nationwide. Unadjusted rate ratios were calculated to compare the relative rates of hospitalization by demographic subgroup or presence of each underlying medical condition. The numerator for each rate was the number of hospitalized adults in each demographic subgroup with or without each underlying medical condition estimated from COVID-NET. The denominator for each rate was the number of adults in each demographic subgroup with or without each underlying medical condition derived from BRFSS estimates for the COVID-NET catchment area. Generalized Poisson regression models with a scaled deviance term to account for overdispersion were used to calculate unadjusted and adjusted rate ratios and 95% confidence intervals (CIs) associated with hospitalization. Multivariable models included an individual underlying medical condition, age, sex, and race/ethnicity. Model goodness of fit was assessed by evaluating standardized deviance residual plots. Rate ratios with 95% CIs that excluded 1 were considered statistically significant. We also assessed the prevalence of co-occurring conditions in hospitalized cases (Supplemental Table 2); however, due to the analytic design of this study and small cell counts of BRFSS estimates from the COVID-NET catchment area, we were unable to account for combinations of underlying medical conditions in our adjusted models. Weighted population estimates from BRFSS were calculated using SAS-callable SUDAAN. All other analyses were performed using SAS v.9.4 (SAS Institute, Cary, NC).

No personal identifiers were included in either COVID-NET or BRFSS data submitted to CDC. This analysis was exempt from CDC’s Institutional Review Board, as it was considered part of public health surveillance and emergency response. Participating sites obtained approval for the COVID-NET surveillance protocol from their respective state and local IRBs, as required.

## RESULTS

Of 5,416 community-dwelling adults with COVID-19-associated hospitalization, 30% were aged 18-44 years, 40% were aged 45-64 years and 31% were aged 65+ years; 53% were male; 34% were non-Hispanic White, 32% were non-Hispanic Black and 34% were of other races/ethnicities (Table 1). Overall, 55% had obesity, 49% had hypertension, 33% had diabetes, 16% had severe obesity, 13% had asthma, 12% had chronic kidney disease, 9% had a history of coronary artery disease, 6% had COPD, and 4% had a history of stroke. Excluding hypertension, 73% of hospitalized cases had at least one underlying medical condition. Co-occurring underlying medical conditions were common among hospitalized cases (e.g., most adults with coronary artery disease, stroke, diabetes, chronic kidney disease, or COPD also had hypertension) (Supplemental Table 2).

**Table 1:**
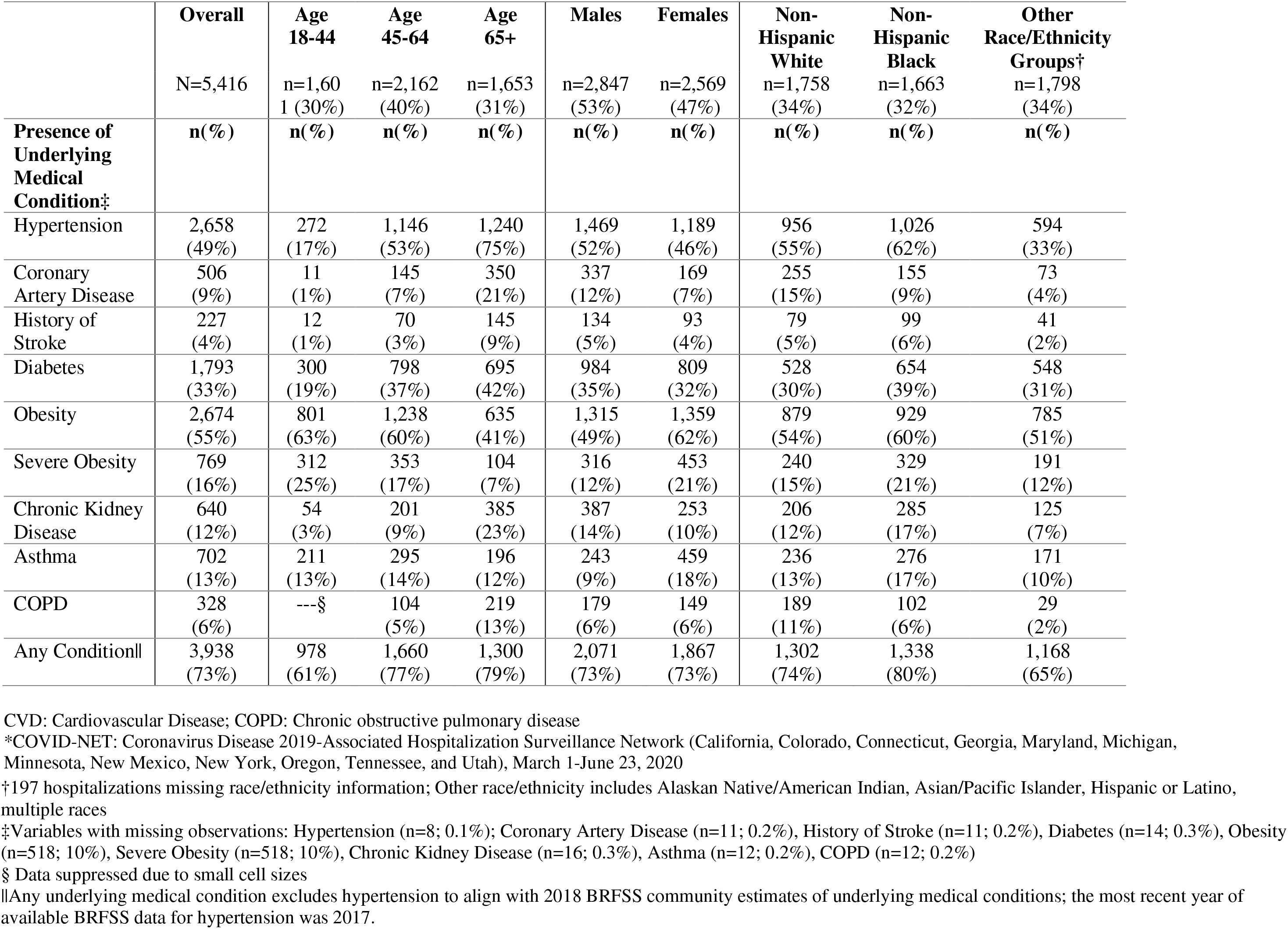
Prevalence of Specific Underlying Medical Conditions among Community Dwelling Adults with COVID-19-associated Hospitalizations by Age, Sex, and Race/Ethnicity, COVID-NET* (N=5,416)

|  | **Overall** | **Age**<br>**18-44** | **Age**<br>**45-64** | **Age**<br>**65+** | **Males** | **Females** | **Non-**<br>**Hispanic**<br>**White** | **Non-**<br>**Hispanic**<br>**Black** | **Other**<br>**Race/Ethnicity**<br>**Groups†** |
| --- | --- | --- | --- | --- | --- | --- | --- | --- | --- |
|  | N=5,416 | n=1,601<br>(30%) | n=2,162<br>(40%) | n=1,653<br>(31%) | n=2,847<br>(53%) | n=2,569<br>(47%) | n=1,758<br>(34%) | n=1,663<br>(32%) | n=1,798<br>(34%) |
| **Presence of**<br>**Underlying**<br>**Medical**<br>**Condition‡** | **n(%)** | **n(%)** | **n(%)** | **n(%)** | **n(%)** | **n(%)** | **n(%)** | **n(%)** | **n(%)** |
| Hypertension | 2,658<br>(49%) | 272<br>(17%) | 1,146<br>(53%) | 1,240<br>(75%) | 1,469<br>(52%) | 1,189<br>(46%) | 956<br>(55%) | 1,026<br>(62%) | 594<br>(33%) |
| Coronary<br>Artery Disease | 506<br>(9%) | 11<br>(1%) | 145<br>(7%) | 350<br>(21%) | 337<br>(12%) | 169<br>(7%) | 255<br>(15%) | 155<br>(9%) | 73<br>(4%) |
| History of<br>Stroke | 227<br>(4%) | 12<br>(1%) | 70<br>(3%) | 145<br>(9%) | 134<br>(5%) | 93<br>(4%) | 79<br>(5%) | 99<br>(6%) | 41<br>(2%) |
| Diabetes | 1,793<br>(33%) | 300<br>(19%) | 798<br>(37%) | 695<br>(42%) | 984<br>(35%) | 809<br>(32%) | 528<br>(30%) | 654<br>(39%) | 548<br>(31%) |
| Obesity | 2,674<br>(55%) | 801<br>(63%) | 1,238<br>(60%) | 635<br>(41%) | 1,315<br>(49%) | 1,359<br>(62%) | 879<br>(54%) | 929<br>(60%) | 785<br>(51%) |
| Severe Obesity | 769<br>(16%) | 312<br>(25%) | 353<br>(17%) | 104<br>(7%) | 316<br>(12%) | 453<br>(21%) | 240<br>(15%) | 329<br>(21%) | 191<br>(12%) |
| Chronic Kidney<br>Disease | 640<br>(12%) | 54<br>(3%) | 201<br>(9%) | 385<br>(23%) | 387<br>(14%) | 253<br>(10%) | 206<br>(12%) | 285<br>(17%) | 125<br>(7%) |
| Asthma | 702<br>(13%) | 211<br>(13%) | 295<br>(14%) | 196<br>(12%) | 243<br>(9%) | 459<br>(18%) | 236<br>(13%) | 276<br>(17%) | 171<br>(10%) |
| COPD | 328<br>(6%) | ---§ | 104<br>(5%) | 219<br>(13%) | 179<br>(6%) | 149<br>(6%) | 189<br>(11%) | 102<br>(6%) | 29<br>(2%) |
| Any Condition\|\| | 3,938<br>(73%) | 978<br>(61%) | 1,660<br>(77%) | 1,300<br>(79%) | 2,071<br>(73%) | 1,867<br>(73%) | 1,302<br>(74%) | 1,338<br>(80%) | 1,168<br>(65%) |

CVD: Cardiovascular Disease; COPD: Chronic obstructive pulmonary disease
\*COVID-NET: Coronavirus Disease 2019-Associated Hospitalization Surveillance Network (California, Colorado, Connecticut, Georgia, Maryland, Michigan, Minnesota, New Mexico, New York, Oregon, Tennessee, and Utah), March 1-June 23, 2020
† 197 hospitalizations missing race/ethnicity information; Other race/ethnicity includes Alaskan Native/American Indian, Asian/Pacific Islander, Hispanic or Latino, multiple races
‡ Variables with missing observations: Hypertension (n=8; 0.1%); Coronary Artery Disease (n=11; 0.2%), History of Stroke (n=11; 0.2%), Diabetes (n=14; 0.3%), Obesity (n=518; 10%), Severe Obesity (n=518; 10%), Chronic Kidney Disease (n=16; 0.3%), Asthma (n=12; 0.2%), COPD (n=12; 0.2%)
§ Data suppressed due to small cell sizes
|| Any underlying medical condition excludes hypertension to align with 2018 BRFSS community estimates of underlying medical conditions; the most recent year of available BRFSS data for hypertension was 2017.

Among hospitalized cases, the prevalence of underlying medical conditions was greatest among adults aged 65+ years except for obesity, severe obesity, and asthma (Table 1). The prevalence of obesity (63%) and severe obesity (25%) was greatest among adults aged 18-44 years. The prevalence of asthma was similar across all age groups. Males and females had similar prevalences of history of stroke, diabetes, and COPD. The prevalence of underlying medical conditions was highest among non-Hispanic black adults, except for coronary artery disease and COPD.

The overall prevalence of selected underlying medical conditions was greater among hospitalized cases compared to the COVID-NET catchment area population (Figure 2). COVID-NET catchment area estimates were similar or slightly lower than nationwide estimates: hypertension (21% vs. 25%), coronary artery disease (5% vs. 7%), history of stroke (3% vs. 3%), diabetes (9% vs. 11%), obesity (28% vs. 31%), severe obesity (4% vs 5%), chronic kidney disease (2% vs. 3%), asthma (10% vs. 9%), and COPD (5% vs. 7%).

**Figure 2.**
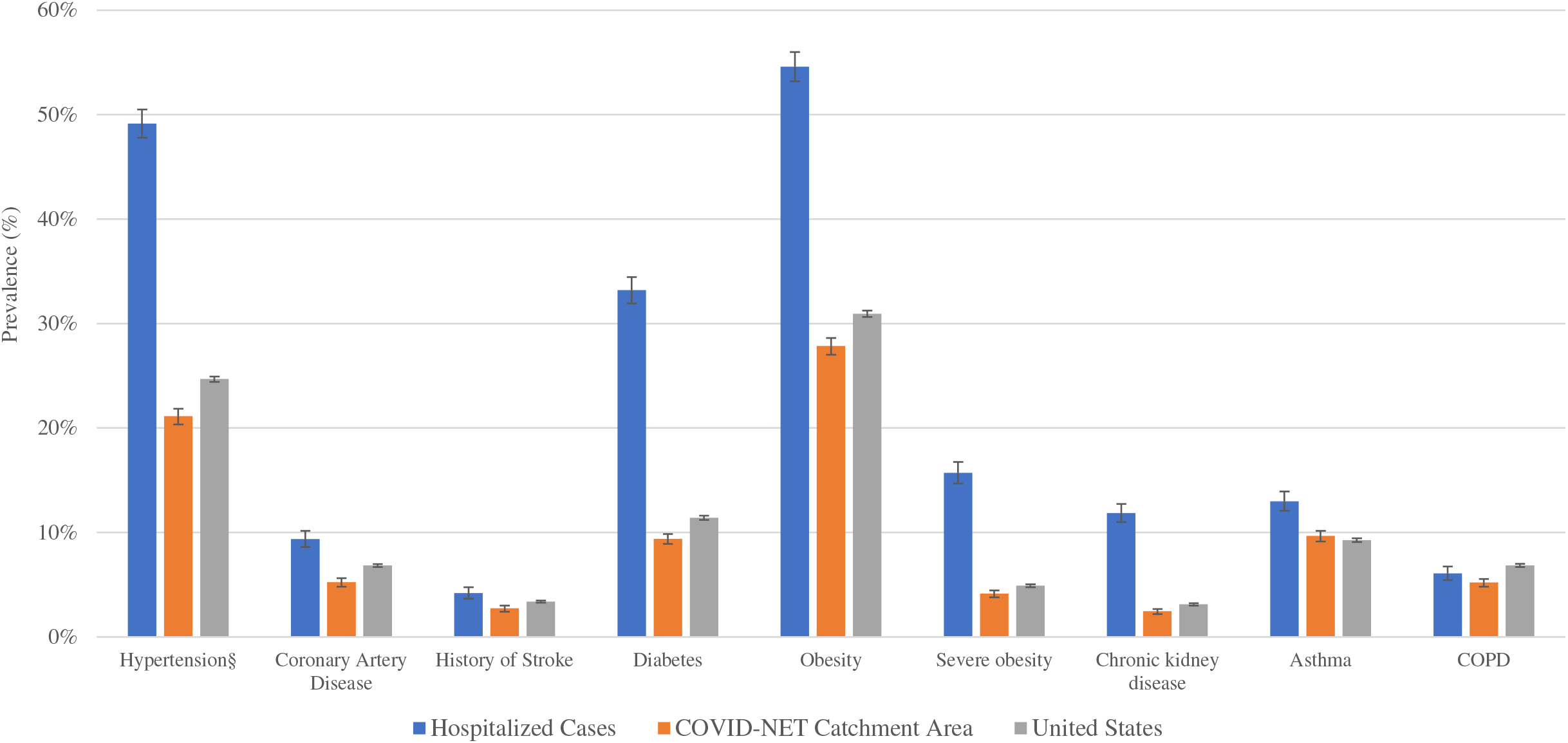
Prevalence of Underlying Medical Conditions: Community Dwelling Adults with COVID-19-associated Hospitalizations,* COVID-NET Catchment Population,† and Nationwide BRFSS Estimates‡. COPD: Chronic obstructive pulmonary disease *Prevalence of underlying medical conditions among community-dwelling hospitalized cases from COVID-NET: Coronavirus Disease 2019-Associated Hospitalization Surveillance Network (COVID-NET), March 1-June 23, 2020; error bars represent 95% confidence interval surrounding estimates †Catchment population estimates from direct Behavioral Risk Factor Surveillance System estimates of underlying medical conditions aggregated from counties participating in COVID-NET, providing community level data on underlying health conditions, 2018; error bars represent 95% confidence interval surrounding estimates ‡Nationwide estimates from Behavioral Risk Factor Surveillance System (BRFSS), 2018; error bars represent 95% confidence interval surrounding estimates §Estimates for hypertension from COVID-NET Catchment Area and Nationwide BRFSS estimates are from 2017, the latest year of available data.

Unadjusted rate ratios for COVID-19-associated hospitalizations of adults 45-64 years of age and 65 years and older, versus 18-44 years, were 2.0 (95%CI: 1.8, 2.1) and 2.7 (95%CI: 2.5, 2.9), respectively (Table 2). The unadjusted rate ratio for hospitalization comparing males to females was 1.2 (95%CI: 1.1, 1.3) and for non-Hispanic black to non-Hispanic white adults was 3.9 (95%CI: 3.7, 4.2). Adults with, versus without, specified underlying medical conditions had higher rates of hospitalization; unadjusted rate ratios ranged from 1.2 (95%CI: 0.4, 3.8) for COPD to 5.3 (95%CI: 2.4, 12.1) for chronic kidney disease.

**Table 2.**
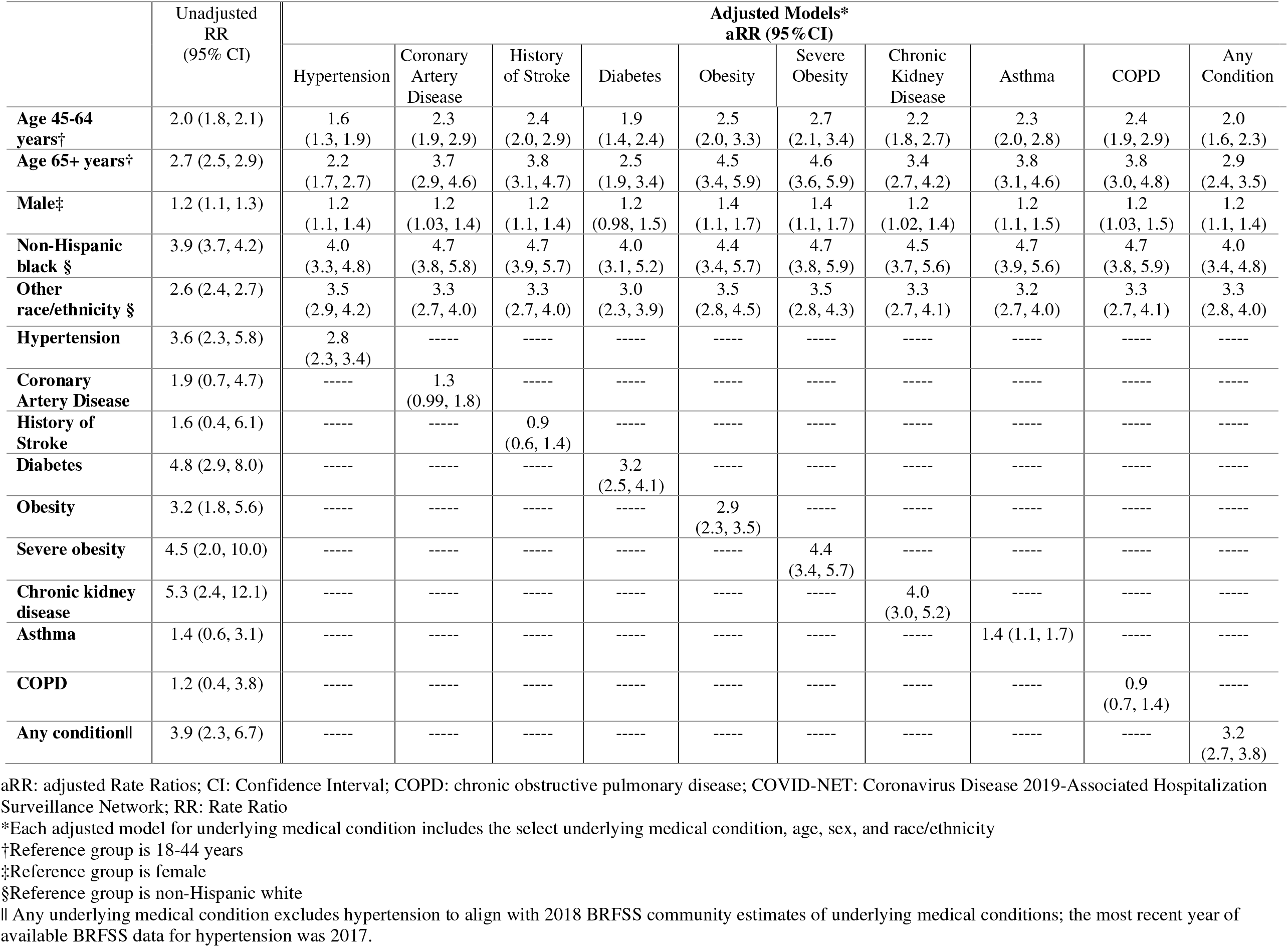
Unadjusted and Adjusted* Rate Ratios for COVID-19-Associated Hospitalizations by Underlying Condition among Community Dwelling Adults, COVID-NET, March 1-June 23, 2020.

The rate ratios for underlying medical conditions attenuated after adjustment for age, sex, and race/ethnicity; except for the rate ratios for severe obesity and asthma which remained stable (Table 2). In descending order of magnitude, the adjusted rate ratios (aRR) for hospitalization by underlying medical condition were as follows: severe obesity (aRR:4.4; 95%CI: 3.4, 5.7), chronic kidney disease (aRR:4.0; 95%CI: 3.0, 5.2), diabetes (aRR:3.2; 95%CI: 2.5, 4.1), obesity (aRR:2.9; 95%CI: 2.3, 3.5), hypertension (aRR:2.8; 95%CI: 2.3, 3.4), asthma (aRR:1.4; 95%CI: 1.1, 1.7), coronary artery disease (aRR:1.3; 95%CI:0.99, 1.8), COPD (aRR: 0.9; 95%CI: 0.7, 1.4), stroke (aRR: 0.9; 95%CI: 0.6, 1.4) (Table 2; Figure 3). After adjustment for age, sex, and race/ethnicity, rate ratios for hospitalization increased with the number of conditions (versus no conditions), with the greatest rate ratio for adults with 3+ conditions (aRR: 5.0; 95%CI:3.9, 6.3) (Supplemental Table 3).

**Figure 3:**
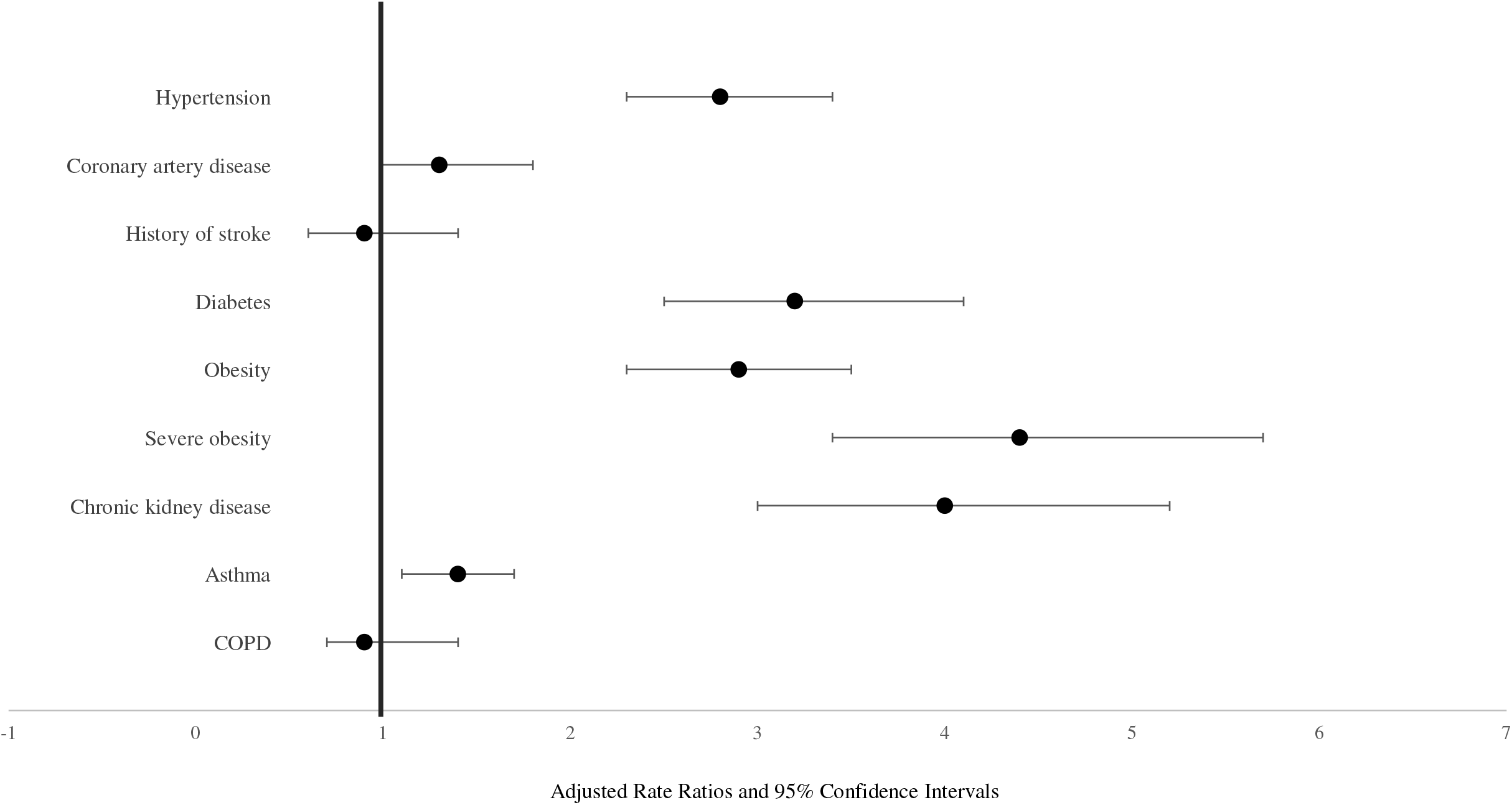
Adjusted* Rate Ratios for COVID-19-Associated Hospitalization by Underlying Medical Condition, COVID-NET, March 1-June 23, 2020. COPD: Chronic obstructive pulmonary disease; COVID-NET: Coronavirus Disease 2019-Associated Hospitalization Surveillance Network *Adjusted for age, sex, race/ethnicity; also shown in Table 2

Across individual underlying medical condition models, the adjusted rate ratio of hospitalization was significantly higher for adults ≥65 years and 45-64 years (versus 18-44 years), males (versus females), and non-Hispanic black and other race/ethnicities (versus non-Hispanic whites) (Table 2). For example, in the severe obesity model, adults ≥65 years (aRR: 4.6; 95%CI: 3.6, 5.9), 45-64 years (aRR: 2.7; 95%CI 2.1, 3.4) versus18-44 years; males versus females (aRR: 1.4; 95%CI: 1.1, 1.7); and non-Hispanic blacks (aRR: 4.7; 95%CI: 3.8, 5.9) and other race/ethnicities (aRR: 3.5; 95%: 2.8, 4.3) versus non-Hispanic whites had higher hospitalization rates. These associations were similar in models adjusting for any condition (Table 2) and number of conditions (Supplemental Table 3).

## DISCUSSION

In this study utilizing two large multi-state surveillance systems to compare hospitalized cases with the community at risk, we found that increasing age, male sex, non-Hispanic black race/ethnicity, other race/ethnicities, and select underlying medical conditions were associated with a significantly greater risk for COVID-19-associated hospitalization relative to the non-hospitalized community-dwelling adult population. Among the underlying medical conditions studied, the magnitude of risk was greatest for severe obesity, chronic kidney disease, diabetes, obesity, and hypertension; each of these conditions was independently associated with approximately 3 or more times the risk of hospitalization after accounting for age, sex, and race/ethnicity. Among adults who tested positive for SARS-CoV-2 and sought care at health systems in Louisiana (19) and in New York City and Long Island (20), chronic kidney disease (20), obesity (19, 20), diabetes (20), and hypertension (20) were also found to be associated with increased odds of hospitalization (adjusted odds ratios ranging from 1.4 to 2.4) after accounting for age, sex, race/ethnicity (19, 20), and either the Charlson comorbidity index (19) or other select medical conditions (20). Our study extends the literature by quantifying the independent association of underlying medical conditions with hospitalization relative to the community population at risk.

Similar to other studies (10, 11, 12, 13, 14), we found that hypertension, obesity, and diabetes were common among COVID-19-associated hospitalizations. In our study, prevalences of all select underlying medical conditions was greatest among hospitalized COVID-19 patients compared to the COVID-NET catchment area and nationwide. Similar to nationwide estimates (16, 17), hypertension and diabetes were more common in middle and older aged adults with COVID-19-associated hospitalizations. Obesity was greatest in those 18-44 years old with COVID-19-associated hospitalizations, unlike nationwide estimates of obesity which are relatively similar across age groups (15). The prevalence of chronic kidney disease among adults with COVID-19-associated hospitalizations was similar to national estimates of chronic kidney disease calculated from albuminuria or serum creatinine measures (23). However, these estimates were higher than self-reported estimates from the COVID-NET catchment area and nationwide derived estimates from BRFSS. This difference may be in part due to medical abstraction vs. self-report ascertainment; an estimated 90% of adults with chronic kidney disease do not know they have it (23).

The magnitude of risk for COVID-19-associated hospitalization was lower for coronary artery disease, stroke, asthma, and COPD than for other medical conditions (e.g., hypertension) in our analysis. Our prevalence estimates of asthma and/or COPD (18%) was similar to a study among adults who tested positive for SARS-CoV-2 (15%), which found that asthma or COPD was not independently associated with risk for hospitalization (20). However, among hospitalized patients, coronary artery disease and COPD have both been found to be associated with intensive care unit admission, need for mechanical ventilation (24,25) and mortality (24, 25, 26).

We found that ages 45-64 years and 65+ years were independently associated with increased risk of hospitalization compared to ages 18-44 years after accounting for underlying medical conditions, sex, and race/ethnicity. Further, the magnitude of risk for hospitalization was greatest among adults 65 years and older, similar to other studies (19, 20). It is important to note that the additional risk of age 65 years and older, and of age 45-64 years, is relative to younger age (18-44 years) and should not be interpreted as absolute risk.

Males were 30% more likely to be hospitalized than females after accounting for age, race/ethnicity, and underlying medical conditions, similar to another study (20). Non-biological factors may lead to a greater proportion of males being hospitalized (e.g., increased exposure or delays in care seeking). Biological factors could include immune function suppression by testosterone compared to estrogen (27) or lower expression of angiotensin-converting enzyme 2, a receptor that allows entry of SARS-CoV-2 into host cells, due to estrogen, potentially inhibiting severe clinical progression in females compared to males (28).

Over-representation of non-Hispanic black adults among hospitalized COVID-19 patients has been hypothesized to be due to the higher prevalence of underlying medical conditions (10, 19) such as hypertension, obesity, diabetes, and chronic kidney disease among the non-Hispanic black population (15, 16, 17, 23). While these conditions contributed to the total risk, we found that after accounting for underlying medical conditions, age, and sex, non-Hispanic black adults had four times greater risk of hospitalization than non-Hispanic white adults. Additionally, the magnitude of risk was similar across underlying medical conditions (aRR range: 4.0 to 4.7), suggesting that non-Hispanic black adults experience excess risk regardless of select underlying medical conditions. This association was also observed when controlling for the presence of any condition or the number of conditions. Black race was similarly associated with 3 times the risk of hospitalization in an Atlanta-based study (29). It has been suggested that non-Hispanic black adults might be more likely to be hospitalized due to increased exposures (e.g., related to occupation or housing) that could lead to increased incidence or more severe illness; differences in health care access or utilization; or systemic social inequities, including racism and discrimination (30, 31, 32). However, we were unable to assess these factors with our data. These factors may also explain similar findings of increased risk for hospitalization among other race/ethnicities.

Overall, these results have implications for clinical practice, as they identify high-risk patients who require closer monitoring and management of their chronic conditions during the ongoing COVID-19 pandemic. While specific underlying medical conditions studied imparted higher risk of hospitalization, we were unable to account for the duration of each condition or the degree to which each condition was controlled (e.g., glycemic control in diabetic patients). Nevertheless, clinicians might prioritize more aggressive control of underlying conditions with available treatments and encourage their patients to remain engaged in care for management of their chronic conditions while practicing preventive measures, such as wearing a cloth face covering and social distancing. These groups may also benefit from targeted preventative and therapeutic interventions.

### Limitations

This study has several limitations. First, this analysis is based on data as of June 23, 2020 from COVID-NET, a surveillance system designed first to provide hospitalization rates. Additional data such as underlying medical conditions is reliant on medical chart abstraction; approximately 60% of the total hospitalized cases have yet to be abstracted for underlying medical condition. Thus, included cases represent a convenience sample of hospitalizations with underlying medical conditions, which may have resulted in biased estimates of risk. However, bi-weekly updates of this analysis over a 2-month period with the most recently available COVID-NET data (i.e., additional chart abstractions) suggests consistent estimates of the frequency and distribution of underlying conditions and resulting rate ratios. Second, these data did not include institutionalized adults. Third, estimates of risk are restricted to the COVID-NET catchment area; the interpretation of rate ratios as risk in this analysis assumes that risk of SARS-CoV-2 infection is consistent across all groups. Fourth, we were unable to assess the association of more granular race/ethnicity categories or co-occurring underlying health conditions due to small cell sizes from the COVID-NET catchment area; further investigation on both aspects is important. Fifth, COVID-NET likely under-ascertains COVID-19 cases as testing for SARS-CoV-2 is performed at treating health care providers’ discretion and is subject to clinician bias as well as variability in testing practices and capabilities across providers and facilities. However, this probably had minimal impact on our findings as hospitalized individuals are more likely to be tested than those in the community. Finally, we used BRFSS to obtain estimates for underlying medical conditions in the COVID-NET catchment area. As the ascertainment of underlying medical conditions was different across the two data systems (self-report vs. medical chart abstraction), we may have introduced bias in the rate ratio estimation. Self-reported diabetes (33) and hypertension (34) have high correlation with medical examination estimates. Self-report has been found to underestimate prevalence of chronic kidney disease (23) and obesity (34); thus, our rate ratios for these conditions may be overestimated.

## CONCLUSION

This analysis quantifies associations of age, sex, race/ethnicity, and underlying medical conditions with risk of COVID-19 hospitalization relative to the non-hospitalized community-dwelling population. These data may aid clinicians in identifying individuals at higher risk for hospitalization who may require more vigilant care and monitoring, and groups that may benefit from preventive and therapeutic interventions.

## Data Availability

Data is not publically available at this time.

## ACKNOWLEDGEMENTS

Erin Parker, Jeremy Roland, Gretchen Rothrock (California Emerging Infections Program); Isaac Armistead, Rachel Herlihy, Sarah McLafferty (Colorado Department of Public Health and Environment); Paula Clogher, Hazal Kayalioglu, Amber Maslar, Adam Misiorski, Linda Niccolai, Danyel Olson, Christina Parisi (Connecticut Emerging Infections Program, Yale School of Public Health); Emily Fawcett, Katelyn Lengacher, Jeremiah Williams (Emerging Infections Program, Georgia Department of Health, Veterans Affairs Medical Center, Foundation for Atlanta Veterans Education and Research); Jim Collins, Kimberly Fox, Sam Hawkins, Shannon Johnson, Libby Reeg, Val Tellez Nunez (Michigan Department of Health and Human Services); Erica Bye, Richard Danila, Nagi Salem (Minnesota Department of Health); Kathy Angeles, Lisa Butler, Cory Cline, Kristina G. Flores, Caroline Habrun, Emily B. Hancock, Sarah Khanlian, Meaghan Novi, Erin C. Phipps (New Mexico Emerging Infections Program); Alison Muse, Adam Rowe (New York State Department of Health); Sophrena Bushey, Maria Gaitan, RaeAnne Kurtz, Marissa Tracy (Rochester Emerging Infections Program, University of Rochester Medical Center); Ama Owusu-Dommey, Lindsey Snyder (Oregon Health Authority); Katherine Michaelis, Kylie Seeley (Oregon Health & Science University School of Medicine); Kathy Billings, Katie Dyer, Melinda Eady, Anise Elie, Gaily Hughett, Karen Leib, Tiffanie Markus, Terri McMinn, Danielle Ndi, Manideepthi Pemmaraju, John Ujwok (Vanderbilt University Medical Center); Ryan Chatelain, Andrea George, Keegan McCaffrey, Jacob Ortega, Andrea Price, Ilene Risk, Melanie Spencer, Ashley Swain (Salt Lake County Health Department); Rainy Henry, Sonja Nti-Berko, Bob Pinner, Alvin Shultz (Emerging Infections Program); Mimi Huynh, Monica Schroeder (Council for State and Territorial Epidemiologists); Junling Ren, Bill Bartoli, Liegi Hu (CDC Division of Population Health, Northrup Grumman); Gayle Langley, Melissa Rolfes, Carrie Reed (CDC).

## Notes

### Competing Interest Statement

Dr. Anderson reports personal fees from AbbVie, personal fees from Pfizer, grants from Pfizer, grants from Merck, grants from Micron, grants from Paxvax, grants from Sanofi Pasteur, grants from Novavax, grants from MedImmune, grants from Regeneron, grants from GSK, outside the submitted work. Mr. Henderson, Ms. Kim, Ms. George, and Ms. Hill report grants from Council of State and Territorial Epidemiologists (CSTE), during the conduct of the study. Dr. Lynfield reports grants from CDC- Emerging Infections Program, during the conduct of the study; and Royalties from a book on infectious disease surveillance and compensation for AAP Red Book (Report from Committee on Infectious Disease) donated to Minnesota Dept of Health. Dr. Schaffner reports grants from CDC, during the conduct of the study; personal fees from VBI Vaccines, outside the submitted work. Dr. Talbot reports other from Seqirus, outside the submitted work.

### Funding Statement

This work was supported by the Centers of Disease Control and Prevention through an Emerging Infections Program cooperative agreement (grant CK17-1701) and through a Council of State and Territorial Epidemiologists cooperative agreement (grant NU38OT000297-02-00).

### Author Declarations

This analysis was exempt from CDC's Institutional Review Board, as it was considered part of public health surveillance and emergency response. Participating sites obtained approval for the COVID-NET surveillance protocol from their respective state and local IRBs, as required.

